# Forecasting hospital-level COVID-19 admissions using real-time mobility data

**DOI:** 10.1101/2022.06.06.22275840

**Authors:** Brennan Klein, Ana C. Zenteno, Daisha Joseph, Mohammadmehdi Zahedi, Michael Hu, Martin Copenhaver, Moritz U.G. Kraemer, Matteo Chinazzi, Michael Klompas, Alessandro Vespignani, Samuel V. Scarpino, Hojjat Salmasian

## Abstract

For each of the COVID-19 pandemic waves, hospitals have had to plan for deploying surge capacity and resources to manage large but transient increases in COVID-19 admissions. While a lot of effort has gone into predicting regional trends in COVID-19 cases and hospitalizations, there are far fewer successful tools for creating accurate hospital-level forecasts. At the same time, anonymized phone-collected mobility data proved to correlate well with the number of cases for the first two waves of the pandemic (spring 2020, and fall-winter 2021). In this work, we show how mobility data could bolster hospital-specific COVID-19 admission forecasts for five hospitals in Massachusetts during the initial COVID-19 surge. The high predictive capability of the model was achieved by combining anonymized, aggregated mobile device data about users’ contact patterns, commuting volume, and mobility range with COVID hospitalizations and test-positivity data. We conclude that mobility-informed forecasting models can increase the lead-time of accurate predictions for individual hospitals, giving managers valuable time to strategize how best to allocate resources to manage forthcoming surges.

## 1 Introduction

The COVID-19 pandemic has highlighted the importance of proactively managing hospital resources such as staff, beds, medical equipment, and supplies [1–4]. Hospital administrators have had to balance the impending need for redirecting resources for COVID-19-related care with deferring time-sensitive care of non-COVID patients [2, 5–7]. In the United States, hospitals spent years creating plans to surge inpatient capacity in case of an emergency. Very little work was done, however, on modeling the size and temporal scale of potential surges thus severely limiting hospital’s ability to know when and to what extent to implement their surge plans. The pandemic made clear, however, that administrators have a pressing need for frequently updated predictive models that can help them anticipate the amount, the type, and the timing of the resources that will be needed as they try to provide care during a sustained emergency.

Initial work to predict COVID-19 hospital admissions largely focused on using epidemiological models that describe the evolution of a disease within a susceptible population to extrapolate the expected volume of admissions over time [4, 8–11]. These models are most useful for creating a range of long-term scenarios for planners at a city or state-levels who are trying to assess the impact of different mitigation measures. However, it is not clear how to map these projections to individual hospitals. Given their dependency on multiple hardto-estimate parameters such as local community transmission rates, these forecasts typically have very wide confidence intervals; this makes them hard to use for operational leaders of hospitals, who need to make shorter-term, more granular resource allocation decisions [12]. Moreover, even if the epidemiological forecasts were accurate at a population level, our analysis shows how different hospitals within an urban area may not share the load of admissions evenly in time as they serve different populations with different degrees of exposure and risk of hospitalization.

Other statistical approaches to predicting hospital admissions are patient-specific models and models based on environmental data. Patient-specific models estimate the risk of admission based on clinical and social factors in a community [13] or at the moment of presentation at the hospital (e.g., through the emergency department) [14]. These methods may be of limited use during an emerging infectious disease pandemic, as many of the patients admitted during a pandemic may not have any past medical history at the given hospital to be used for inference, and factors influencing each individual’s likelihood of infection or admission may not be represented in their past medical history at all. Environmental-based models use time-varying predictors such as weather and seasonal factors to predict the daily count of patient presentations in the emergency department and corresponding hospital admissions [15–20]. These models rely on relatively large data sets that span multiple years to establish stable underlying seasonal patterns, which are often unavailable for individual hospitals and/or hospital systems, nor for emerging pandemics.

Large-scale mobility data has been an especially promising tool for incorporating realistic mobility flows into epidemiological models. Through increased smartphone use and advances in techniques for preserving user privacy [21], we have seen an increasing number of studies use this type of large-scale, aggregated data. Mobile phone data has been used to examine the impact of COVID-19 mitigation policies on collective behavior [22–24], quantify broad pandemic-related disruptions in our mobility [25–29], and ultimately, inform epidemiological models during the COVID-19 pandemic [30–34]. These data are often aggregated to a specific spatial resolution (e.g., a single county) and can be used to describe changes to typical mobility patterns, commuting behavior, social contact patterns, and inter-regional transit; changes in these metrics have been found to correlate with infection rate and deaths [27]. In this work, we extend these findings further by incorporating commuting and mobility patterns into a model that predicts COVID-19 admissions at the hospital level.

Specifically, we assess the value of incorporating mobility data into statistical predictions of short-term, daily COVID-19 admission patterns for 5 hospitals of varying sizes within a large network in New England during the first two waves of the pandemic. We do this in two ways: i) we assess whether mobility data increases the accuracy of 1-to-2 week projections purely based on previous admission patterns and ii) we explore whether it can increase the lead-time with which accurate projections can be made. The latter is an important operational consideration for managers. Lead time for opening ICU surge capacity can be as short as 3 days [35]; however, the additional lead-time can allow hospitals to properly identify spaces, design staffing plans, re-deploy equipment, and transition out patients that usually use those spaces [36]. This is especially the case in Academic Medical Centers which have very high ICU occupancy levels. We find that while 7or even 14-day-ahead forecasts using only test-positivity rates and prior admissions data are operationally valuable, the inclusion of anonymized, aggregate mobility data provide accurate 21-day-ahead forecasts at the hospital-level.

## 2 Materials and Methods

### 2.1 Data sources

In addition to time series data of statewide test positivity percentages from the Massachusetts Department of Public Health [37], two other data sources were used in this research: aggregated location data from mobile devices and hospital admission data. Because both data sources were only used in aggregated form, the Institutional Review Board (IRB) determined that the study does not qualify as human subject research and is exempt from IRB approval.

#### 2.1.1 Mobile device data

Aggregated location data from mobile devices has been used in a number of studies to quantify different ways that our mobility, social contacts, commuting patterns, etc. have all changed during the COVID-19 pandemic [21, 30–32, 38]. Here, we use data from Cuebiq Inc.; through their *Data for Good* initiative, Cuebiq provided us with large scale, privacy-enhanced (through up-leveling and aggregation) location data from the mobile devices of millions of users. These are first-party data from opted-in users who agree to anonymously share their GPS location data. Cuebiq provides a list of “personal areas” for users in our dataset, which have been up-leveled to the centroid of the census tract they fall in. These personal areas are proxies for the home and work location of users, as they are the most commonly-visited daytime location and most commonly-visited nighttime location.

From a nationwide sample of over 40 million mobile devices, we generate a panel of users with personal areas in the Boston Combined Statistical Area (CSA ID: 148; BostonWorcester-Providence, MA-RI-NH-CT). In order to reduce the noise in user activity (e.g. turnover in device, deleting apps, etc.), we devised strict criteria for inclusion in the panel. Following [27], we include users in our panel who 1) were active since January 7, 2020, 2) were active for at least 21 days per month between January and June, 2020, 3) generated at least one location ping per hour, on average, and 4) had an average geolocation accuracy of less than 50 meters. This resulted in a panel with over 82,000 mobile devices, selected for their accuracy and stability in the dataset. Lastly, the measures derived from this panel are re-weighted (at the county level) by a statistical correction procedure designed to maximize the representativeness of the panel based on county-level demographic data. For more information on the methods involved in this representativeness correction, see [27].

From the location data of users in our panel, we are able to calculate different aggregated measures of mobility, described below. We report each measure as the percent of its “typical” value, which is defined as the day-of-the-week average between January 16 and February 28, 2020. This controls for weekday/weekend variation in activity, such that a value of 100% indicates typical behavior for that day of the week.

##### Estimating commuting volume

Simply, a commute is defined as a user visiting their “home” personal area and their “work” personal area in a single day. A user’s “home” and “work” personal areas are defined using the most common location visited by a user between 9:00pm-5:00am and 9:00am-5:00pm, respectively. These locations are then mapped onto the centroid of the census tract they are in, obscuring users’ true locations. Because personal areas are up-leveled to the census tract, this is a privacy-preserving way to construct a network of commuter flow between tracts, weighted by the number of commutes. To quantify the total commuting volume for a given region, we sum together the number of commutes for users with “home” personal areas in that region.

##### Estimating individual mobility

We estimate individual mobility through a user’s *radius of gyration* [39]. This measure is especially useful for understanding individual mobility because it captures more than simply a user’s *total* distance traveled; rather, the radius of gyration measures how far on average each visited location is from the center of mass of the different locations visited each day. Specifically, the radius of gyration is

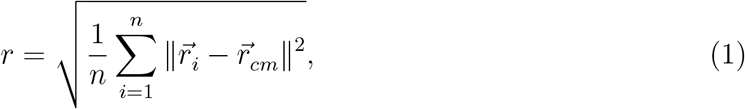

where n is the user’s number of location “pings” on that day, 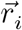 is the i^*th*^ observed location for the user, *i* = 1, 2, …, *n*, and 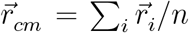 is the center of mass of the trajectory. To quantify the total mobility in a given region, we sum together the daily radii of gyration for users with “home” personal areas in that region.

##### Estimating contact patterns

We also use these data to estimate contact patterns between pairs of users. Because the precise “home” and “work” personal areas of the users in our dataset are obscured to preserve privacy, the contact patterns we measure here are assumed to be contacts outside of home and workplace settings. We compute two measures as proxies for contacts between users—*distinct* contacts per day and *total* contact events per day—as follows. For each location “ping” from a given device, we assign its GPS coordinates to an 8-character *geohash*, which refers to a specific grid cell on the globe. An 8-character geohash is approximately 25m × 25m in size, which may vary based on region on the globe (e.g. 38m × 19m at the equator [40]). For intuitive reference this is slightly larger than a basketball court.

Two users are defined to be co-located if they are observed in the same geohash for at least 15 consecutive minutes—a time frame informed by CDC guidance [41]. For each user, we calculate the total number of contacts with other users as well as the total number of distinct contacts, per day (i.e. if I see one friend three separate times in a day, my number of *distinct* contacts = 1 and my *total* contact events = 3). To quantify the contact patterns for a given region, we average the number of total and distinct contacts of users with “home” personal areas in that region.

#### 2.1.2 Hospitalization data

Admissions data were collected for 10 hospitals from a large non-profit integrated health system in New England. In this system patients have access to a full range of specialty and surgical care, including cancer treatments, neurosurgery, cardiovascular disease, women’s health, and emergency care. For this study, we included hospitalizations from two large academic medical centers, three large community hospitals, and five small community hospitals. We used data from all hospitals for health-system admission computations, and data from the five largest hospitals to make individual site projections; the remaining small community hospitals had very few observations during our time period.

We defined the study period as March 7, 2020 to April 12, 2021. For each calendar day, we count the number of hospital admissions in the system in which patients had an active COVID-19 infection status. A hospitalization is defined as a patient admission where the patient is in an inpatient location (general care floor or intensive care unit) at least once during their admission. The COVID-19 infectious status was considered active if patients had a single confirmed COVID positive Polymerase Chain Reaction (PCR) test during their hospital encounter or if they had presumed COVID due to clinical presentation despite a lack of positive PCR test as determined by the hospitals’ infection control specialists, (typically patients who tested positive by PCR prior to admission). We excluded COVID-19 readmissions, i.e., patients who were hospitalized again within the hospital system with a continuing COVID infection status, because those admissions were not related to the incidence of the disease in the community. We also left out patients who were admitted to the newborn service.

Additionally, we distinguish between “COVID-19 Primary” and “COVID-19 Secondary” admissions. The former correspond to encounters for which COVID was the main reason for admission; they are identified by verifying that the encounters’ admission diagnoses match a list that was manually curated by physicians in our team. The latter correspond to patients who were hospitalized due to reasons different from COVID-19, but had an active COVID-19 infection status during hospitalization (a.k.a. incidental COVID). We make this distinction because we aim to predict COVID-driven admissions on top of what a hospital would have experienced regularly. Primary admissions arrive at the hospital at a specific point of their disease course (5 to 7 days from symptom onset, depending on patient’s age [42–44]) which, we hypothesized, made them more likely to be correlated with mobility patterns as these promote increased interactions in a susceptible population. On the other hand, secondary cases should present to the hospital at a random point of their COVID-19 course given that that’s not their main reason for being admitted. Lastly, if these projections are to be used for resource management, these two groups have very different needs. While they share similar isolation measures, secondary admissions’ care teams correspond to their main reason for admission (e.g., a broken bone), whereas primary admissions will require general medicine care teams, respiratory technicians, proning teams, pulmonologists, ventilators, etc.

Admissions data characteristics are shown in Table 1. It breaks down the COVID-19 hospitalizations during the study period by hospital type and by whether they were COVID-19 Primary or Secondary. The daily counts are aggregated by individual hospital, date of admission, and by the patient’s home ZIP code as captured in the hospital’s electronic medical records (EMR).

**Table 1:** Total number of COVID-19 hospitalizations across the hospitals in our network of study between March 7, 2020 and April 12, 2021. We only considered hospitals A-E for individual site’s projections. Primary and Secondary refer to whether COVID-19 was the main reason for admission. See text for full definitions.

| Hospital ID | Hospital Type | Primary | Secondary | Total |
| --- | --- | --- | --- | --- |
| A | Academic Medical Center | 2,145 | 879 | 3,024 |
| B | Academic Medical Center | 1,257 | 622 | 1,879 |
| C | Large Community Hospital | 1,098 | 407 | 1,505 |
| D | Large Community Hospital | 949 | 282 | 1,231 |
| E | Large Community Hospital | 689 | 229 | 918 |
| F | Small Community Hospital | 238 | 95 | 333 |
| G | Small Community Hospital | 238 | 66 | 304 |
| H | Small Community Hospital | 23 | 7 | 30 |
| I | Small Community Hospital | 17 | 10 | 27 |
| J | Small Community Hospital | 1 | 11 | 12 |
| **Total** |  | **6,655** | **2,608** | **9,263** |

### 2.2 Time series forecasting

Generally, time series forecasting is about predicting the target value at the next step [45]. In the case of COVID-19 hospitalizations, we are working with time series data that are not stationary; that is, the *auto-correlation* between hospital admissions at time step *t* to time step *t* + *k* decreases as k gets larger. Intuitively, this means that knowing today’s number of newly admitted patients is less and less useful for predicting the number of admissions days or weeks from now. To account for this, we treat this task as a supervised learning problem, using previous values in the time series as predictors for the next time step’s value. We do this repeatedly to arrive at a *multi-step forecast*.

For multi-step forecasting, two techniques are commonly used: a direct method and a recursive method [46, 47]. In the direct method, we train a separate model for each forecast time step (e.g. forecasts of *k* = 21 days into the future require 21 separate models). For this approach, there is no information leakage into the future, since each of the time steps from *t* + 1 to *t* + *k* is treated as its own target variable, with input features only extending to time step t. For recursive time series forecasting we first predict the target variable at time step *t* + 1; we then reuse this prediction as an input for the subsequent *t* + 2 time step. This is repeated for k time steps. Here we elected to use this recursive forecasting procedure as it generated more accurate predictions of hospital-level admissions. For recursive forecasting, we used k-step-ahead nonlinear auto regressive forecasting [47] as follows:

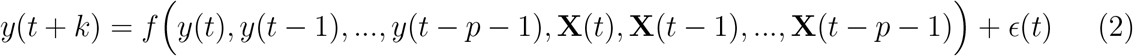

where *y*(*t*) is target time series, **X**(*t*) is multivariate time series of exogenous variables, *p* is the auto regressive order and ϵ is an independent and identically distributed noise term. Function *f*(.) is any regression function. In the following section, we describe the setup, inputs, and training of our recursive forecasts.

#### 2.2.1 Exogenous predictors: Incorporating mobility data

We used daily new COVID-19 Primary admissions as our target forecast and fed an ensemble of exogenous data (based on data from mobile devices, described in Section 2.1.1) directly into the estimator. This kind of exogenous data has been shown to be useful in predicting COVID-19 outcomes throughout the pandemic [11, 27, 30, 34], and here, it makes sense to include as exogenous variables because hospital admissions in this system are due to some extent to the mobility, commuting, and contact patterns of the underlying population; this data is also available in almost real-time, whereas the time between infection, symptoms, and hospitalization for COVID-19 patients can vary by several days.

The measures used in this ensemble are all aggregated to the greater Boston area (i.e., the Boston combined statistical area, or CSA), and they include the following for users in the Cuebiq dataset: average daily commutes per user, average mobility range per user, average number of distinct daily contacts per user, and average number of total daily contacts per user (see Fig. A.2 to how these measures varied throughout the course of a year). Lastly, we also included statewide test positivity time series from the Massachusetts Department of Health [37] as an additional exogenous variable. To generate the final model predictions, we used an XGBoost regression [48] with Poisson loss function; although this technique—“boosting”— is relatively robust against overfitting [49], we added additional L_1_ norm penalties. For each prediction, we generated 95% confidence intervals around the mean based on estimated Poisson-distribution errors.

At the beginning, we took the first four weeks of data, until April 11, 2020 (vertical dashed lines in Fig. 2), for the initial training phase; we split up this initial data into three-week training data and one-week validation data. We trained the model on first portion and tuned hyper-parameters on second portion; then trained the model on the whole first four weeks; then we made forecast for next 21 days; then we added true next day data to training data and re-trained our model on the new training set and made forecasting 21-day ahead and repeat this procedure over and over until we consumed all the data. Selecting a training date this early in the pandemic was deliberate, as it will ultimately highlight the fact that we can generate highly accurate predictions throughout the first year of the pandemic based on a small amount of data.

**Figure 1:**
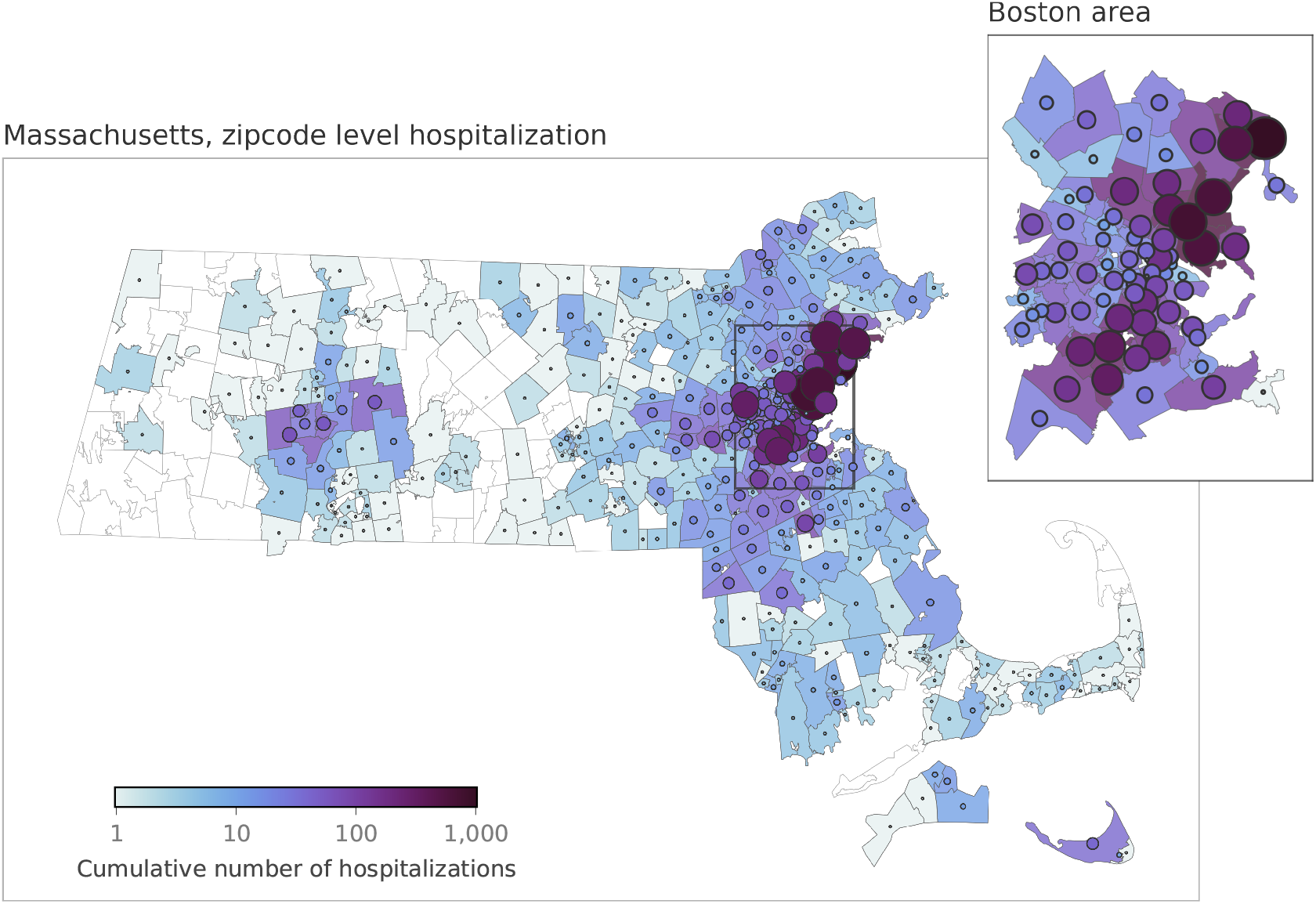
Home address distribution for COVID-19 Primary admissions by ZIP code (Massachusetts only). The five hospitals in this study service a relatively large region around the greater Boston area. Color and size of markers indicate cumulative hospital admissions within ZIP codes.

**Figure 2:**
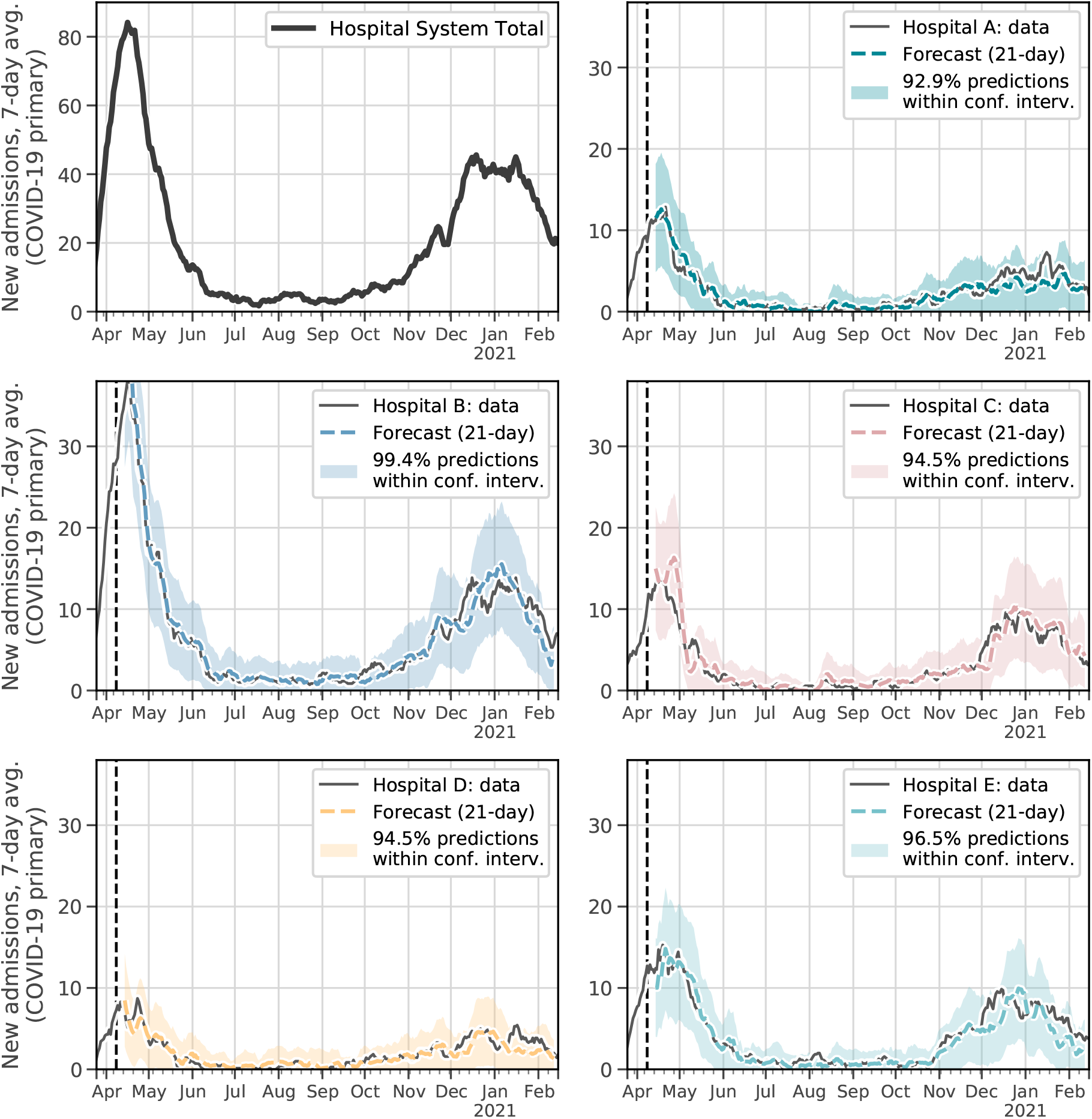
21-day forecast for each hospital. Top left: total hospitalizations across all five hospitals. Each other subplot compares the actual to the predicted hospital admissions, based on the model that incorporates mobility and test positivity data as exogenous variables. The vertical dashed line marks the boundary of boundary of initial training data and future data; visualized at a 7-day rolling average; 95% confidence intervals are shown for each forecast. Hyper-parameters of each model were tuned separately to make sure fair comparisons were made between models.

Ultimately, this approach allows the model to learn (and update, as the pandemic and our behavior co-evolved) associations between the ensemble of exogenous variables and the target variable: new COVID-19 Primary admissions. These forecasts were conducted in Python using the `skforecast` library [50], which transforms the time series data into supervised learning data, i.e. using lagged version of target time series data to predict the current value.

## 3 Results

For each of the five hospitals included in this study, we compared three main forecasts: 1) forecasts that do not include exogenous data (predicting hospitalizations using only previous hospitalizations), 2) forecasts that include mobility data as exogenous variables, 3) forecasts that include Massachusetts-wide test positivity data as exogenous variables, and 4) forecasts that include both mobility and test positivity data as exogenous variables. Ultimately, we show how incorporating aggregated mobile device together with test positivity data into the predictions both increases the overall accuracy of the prediction, and extends this accuracy into a third week of lead time (Fig. 2 and 3). Crucially, the following results are predictions of individual hospitals’ admissions, whereas typically the most effective hospital forecast models are typically performed at the system, county, or state-level.

**Figure 3:**
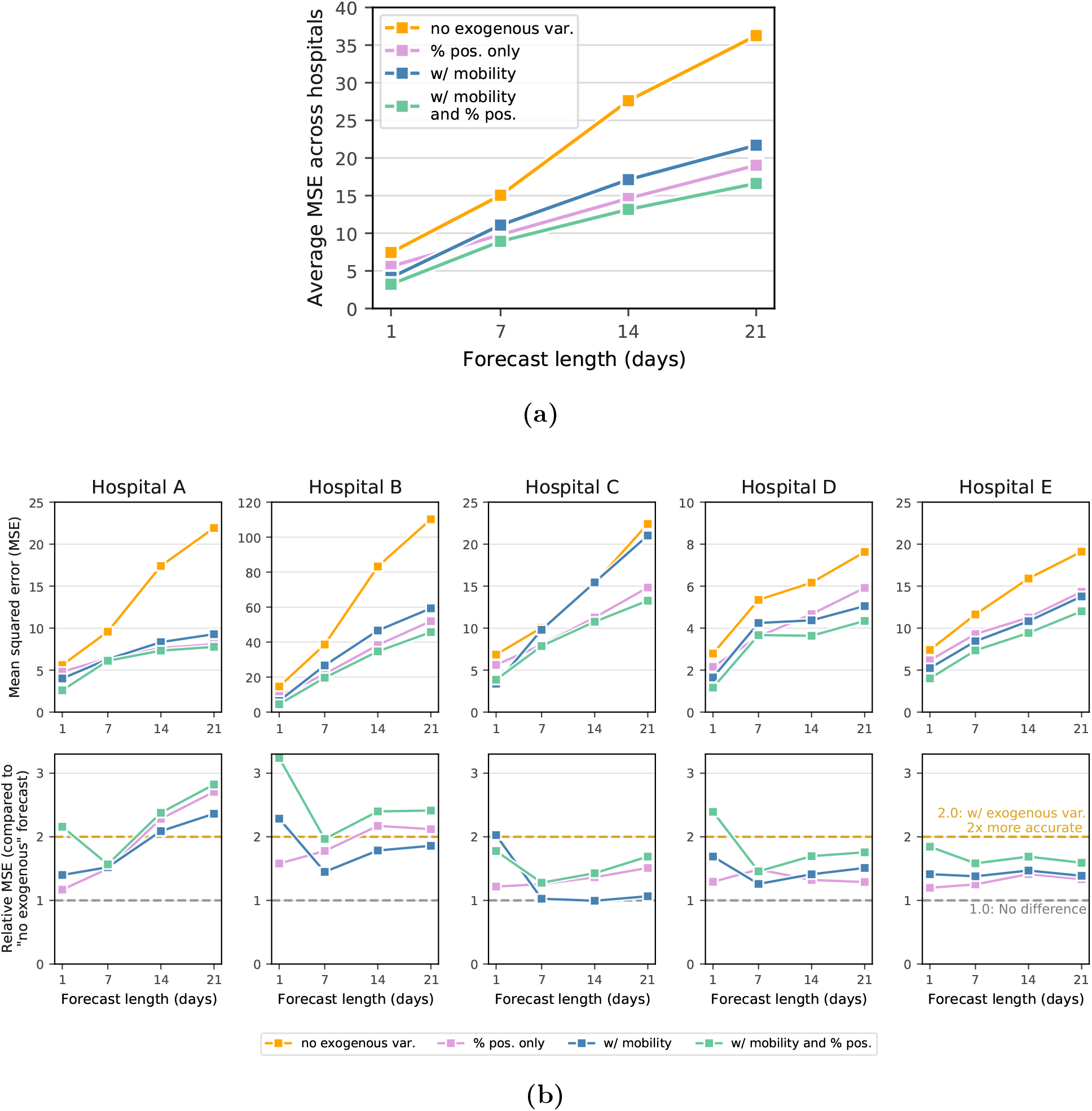
Comparison of forecasts with/without mobility data. **(a)** Average mean squared error (MSE) across all five hospitals under forecasts that 1) do not include exogenous data (orange), 2) include test positivity data from across Massachusetts (pink), 3) include mobility data (blue), and 4) include both mobility and test positivity data (green). **(b)** Top: Hospital-specific MSE, separated by forecast method. Bottom: relative performance, using the forecasts that do not include exogenous variables as a baseline.

Our first observation is perhaps encouraging, especially for hospital systems that do not have access to big data about mobility and social contact patterns: For the hospital system included in this study, we can produce relatively accurate forecasts for system-wide admissions at a 7-day lead time without incorporating additional covariates into the forecast (see Fig. A.3). While 7 days of lead time is useful for planning staffing, room use, and overflow, individual hospitals would be even better served if they could rely on accurate predictions even farther in the future. The accuracy of our 1, 7, 14, and 21-day ahead forecasts improves substantially if we include exogenous data about statewide test positivity (pink lines in Fig. 3).

The core finding in this work is that we generate highly accurate forecasts for hospital admissions 21 days ahead by incorporating exogenous covariates about trends in mobility, commuting, and social contacts, derived from large scale mobile device data (i.e., the only information used by the model was aggregated mobility data described in Section 2.1.1 and previous new admissions). Moreover, these forecasts improve even more after incorporating statewide test positivity as an exogenous variable. In Fig. 2, we overlay time series of the predicted number of hospitalizations for each of the five hospitals in our study with the actual hospital admissions, under the model with these exogenous predictors. By incorporating the mobile device data into our recursive forecasting model, we are able to extend the prediction window more than one week, generating high-accuracy 21-day ahead forecasts at the individual hospital level.

In Fig. 3a, we plot the average mean squared error (MSE) for the entire study period across each of the three models being compared; we do this for different forecast lengths (1, 7, 14, and 21-day forecasts). Under each model, the MSE increases as the length of the forecast increases; this reflects the increasing difficulty of the prediction task as we try to predict farther into the future. However, within each forecast length, we see improved accuracy in the models that do include data about mobility and social contact patterns. We see even higher predictive accuracy when we include test positivity as an exogenous variable. These improvements persist for the 1, 7, 14, and 21-day forecasts, illustrating the power that these kinds of large-scale aggregated mobile device data has in improving our ability to respond to this and future pandemics. As a rough comparison to highlight the accuracy of these forecasts, the MSE of the 21-day forecast that includes both mobility and positivity data is approximately that of the *7-day* forecast that does not incorporate any exogenous predictors and comparable to the *14-day* forecast that only uses positivity data (see Fig. 3a).

Interestingly, among the exogenous (mobility) variables, no single covariate can drive accurate forecasts for the duration of the study period—nor is there a single best-performing combination of covariates throughout the study period. For example, during early Fall 2020, commuting patterns were highly correlated with new admissions (Fig. A.1A), though near the start of 2021, we see that the measures approximating distinct/total daily contacts became more predictive (note, as well, commuting and other mobility measures rarely correlate with *Secondary* COVID-19 admissions—see Fig. A.1B). During the study period, many workplaces started to return in person, restaurants and amenities gradually re-opened [51], and some universities began to bring students back to campus [52]. The societal changes that took place during this period were also reflected in the mobility data—there were several weeks where there was little change in mobility and several weeks when each of the measures varied in its own way. All of this speaks to the dynamic nature of our collective pandemic response and reinforces the need to treat the suite of measures that we can derive from largescale mobility data as an ensemble of collective behavior—a “collective physical distancing” composite measure [27].

## 4 Discussion

We demonstrate that combining aggregated, privacy-preserving mobility data with statewide positivity rates and aggregated hospital admissions data can lead to a more accurate forecast model for future hospital admissions for COVID-19 patients, compared to using hospital admissions data alone. This combined model is particularly accurate—and thus useful for hospital managers—in making a forecast 2-3 weeks in the future. It’s worth mentioning that mobility has its greatest added value in longer-term horizons for the larger community hospitals (Hospitals D and E), which have a smaller number of daily admissions and are likely to be more sensitive to local mobility patterns. Hospital C is different in that it has a very strong relationship with one of the larger Academic Medical Centers and its admissions depend in large part on their partnership.

The association of mobility and COVID-19 transmission is not necessarily causal and certainly subject to various forms of confounding, and the same can be said about the autoregressive relationship between past admissions and future admissions. However, we believe there are signals that make a causal relationship plausible. First, the forecast window of 2-3 weeks is aligned with the typical transmission cycle for COVID-19; that is, despite a serial interval of approximately 5-7 days, 2-3 weeks allows enough time to pick up changes in mobility and contact behavior during non-pharmaceutical interventions. Second, in the latter months of our data, as vaccination efforts started, infection rate and hospitalization rate diminished but mobility trends were increasing at the same time. This new dissociation between mobility data and infection data could suggest that vaccination is breaking a plausibly causal—albeit confounded—relationship between additional exposures that result from higher mobility and the transmission of the disease. Lastly, the fact that these associations are stronger for COVID primary cases compared to COVID secondary cases suggests a plausible causal relationship. That is, the mobile device data used in our model are intended to be proxies for the kinds of interactions that are associated with COVID-19 transmission (e.g. social contacts, co-location for more than 15 minutes, working in-person jobs, etc.).

Our study is not without limitations. First, this study is based on data from one health system (Mass General Brigham) which shares its Health Service Area (HSA) with a few other large and small health systems. The largest hospitals in this health system are inside or close to a major metropolitan area (Boston) and findings from our study may not be representative for other health systems that are not near a metropolitan area or in a shared HSA. One of the health systems within the shared HSA is the Boston Medical Center, which is a safety net hospital with a higher proportion of its patients being from lower socioeconomic groups; this may have also impacted the generalizability of our findings. Second, many of the hospitals in this health system experienced high patient census during the peak of COVID, and patients were transferred between hospitals to help with level loading. This would result in multiple admissions within our system for the same case, and could confound our analysis. While we could not fully account for this due to data quality issues, we do not anticipate this to alter the results. From rough data approximations paired with qualitative statements from bed managers, COVID transfers were not frequent and, if they occurred, we estimate that hospitals mostly admitted 1 to 2 transfers per day.

Our study uses mobility data collected from smart phones; individuals from lower socioeconomic groups may be less likely to own a smartphone or a cellular data plan that would enable continuous use of applications, thereby the amount of mobility data collected for them may be limited, which could impact our findings. Finally, the patterns we saw in the mobility data may be not only influenced by COVID related restrictions (such as lockdown) but also by other secular trends such as seasonality. Because we did not have access to longitudinal pre-COVID mobility data, we could not explore these secular trends.

Anticipating COVID-19 hospital admissions has been a critical feature of capacity management during the pandemic. As hospitals have had to balance scarce resources between COVID and non-COVID patients, this is a very useful tool for clinical operations managers to answer questions such as how much bed capacity is needed and at what pace it has to become available to be able to accommodate a surge in COVID patients. As we showed here, for example, including data about test positivity across the state improves the forecasts even more than including mobility data alone—in particular, comparing the predictions from Fig. 2 and Fig. A.3, this data seems to improve forecasts during periods of low transmission of the virus. This is especially encouraging for hospital administrators between surges in the epidemic, when mobility is less tightly associated with case rates and hospitalizations. Further disentangling the timing and quantity of mobile device data that are needed to achieve these improved forecasts remains an opportunity for future work.

## 5 Conclusion

In this work, we have shown that including high resolution mobility data can help to increase the accuracy of hospital admissions projections with a 3-week lead time. This, in our experience, gives additional precious time to administrators to figure out capacity, staffing, and supplies’ needs. While this type of data is not necessarily readily available to all hospitals, we believe that the benefits of incorporating it are big enough to justify additional effort by city and/or state officials in connecting hospital resources and academic data scientists to make a broader use of this kind of data.

## Data Availability

All data produced in the present study are available upon reasonable request to the authors.

## Additional information

## Acknowledgements

The authors thank Harrison Hartle for helpful feedback on an early draft. Additionally, the authors thank Timothy LaRock, Stefan McCabe, Leo Torres, Maciej Kos, and Lisa Friedland, whose early work was instrumental in defining the mobility measures used in this work.

## Funding information

A.V. and M.C. acknowledge support from COVID Supplement CDC-HHS-6U01IP001137-01 and Cooperative Agreement no. NU38OT000297 from the Council of State and Territorial Epidemiologists (CSTE). M.C. and A.V. acknowledge support from the Google Cloud Research Credits program. A.V. acknowledges support from the McGovern Foundation and the Chleck Family Foundation. The findings and conclusions in this study are those of the authors and do not necessarily represent the official position of the funding agencies, The Rockefeller Foundation, the National Institutes of Health, or U.S. Department of Health and Human Services. The funders had no role in study design, data collection and analysis, decision to publish, or preparation of the manuscript.

## Competing interests

S.V.S. holds unexercised options in Iliad Biotechnologies. This entity provided no financial support associated with this research, did not have a role in the study design, and did not have any role during its execution, analyses, interpretation of the data or decision to submit.

## Data availability

The data underlying this article will be shared on reasonable request to the corresponding author. The Cuebiq Inc. Data for Good initiative is currently known as Spectus and more information can be found at https://spectus.ai/.

## A Supplemental Figures

**Figure A.1:**
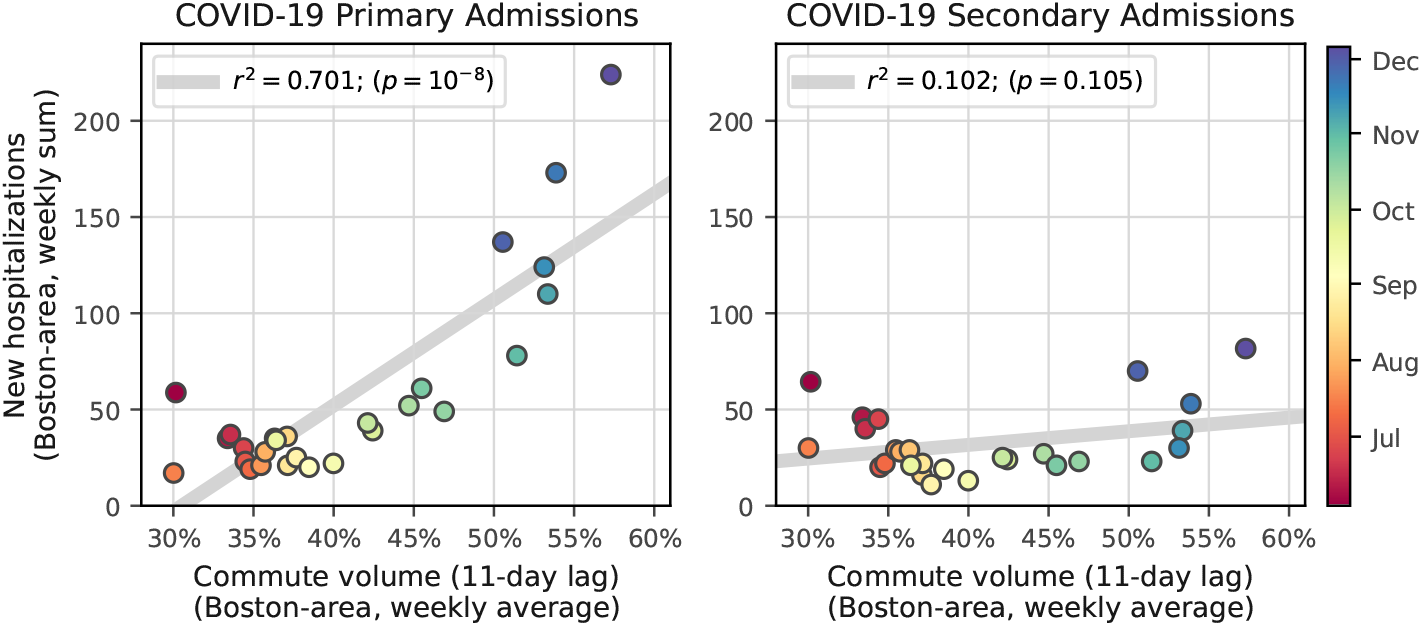
Commute volume and hospital admissions. From June to December 2020, commute volume began to increase in the greater Boston area. This coincided with an increase in weekly COVID-19 “primary” hospital admissions (left), though we do not see the same correlations with COVID-19 “secondary” admissions (right).

**Figure A.2:**
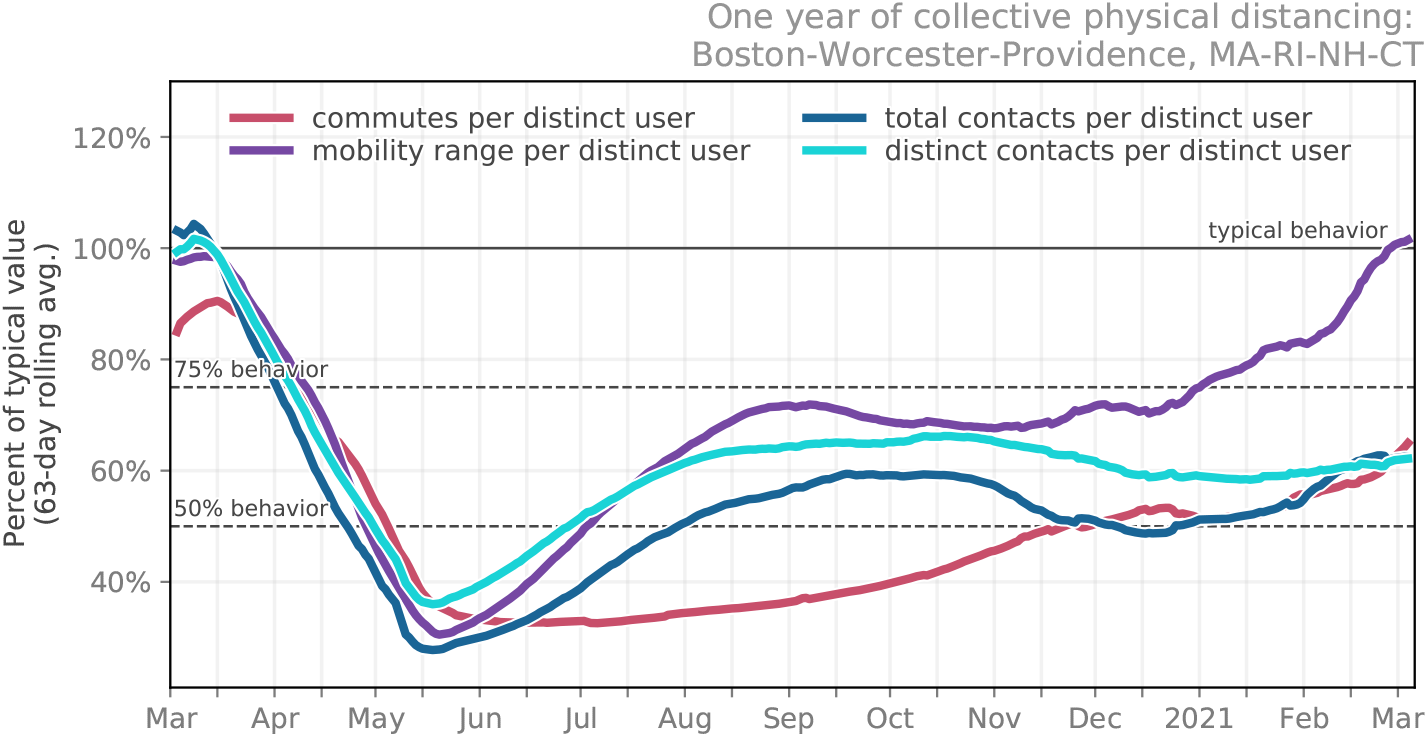
Mobility and contact patterns of the greater Boston combined statistical area. (CSA; Boston-Worcester-Providence, MA-RI-NH-CT).

**Figure A.3:**
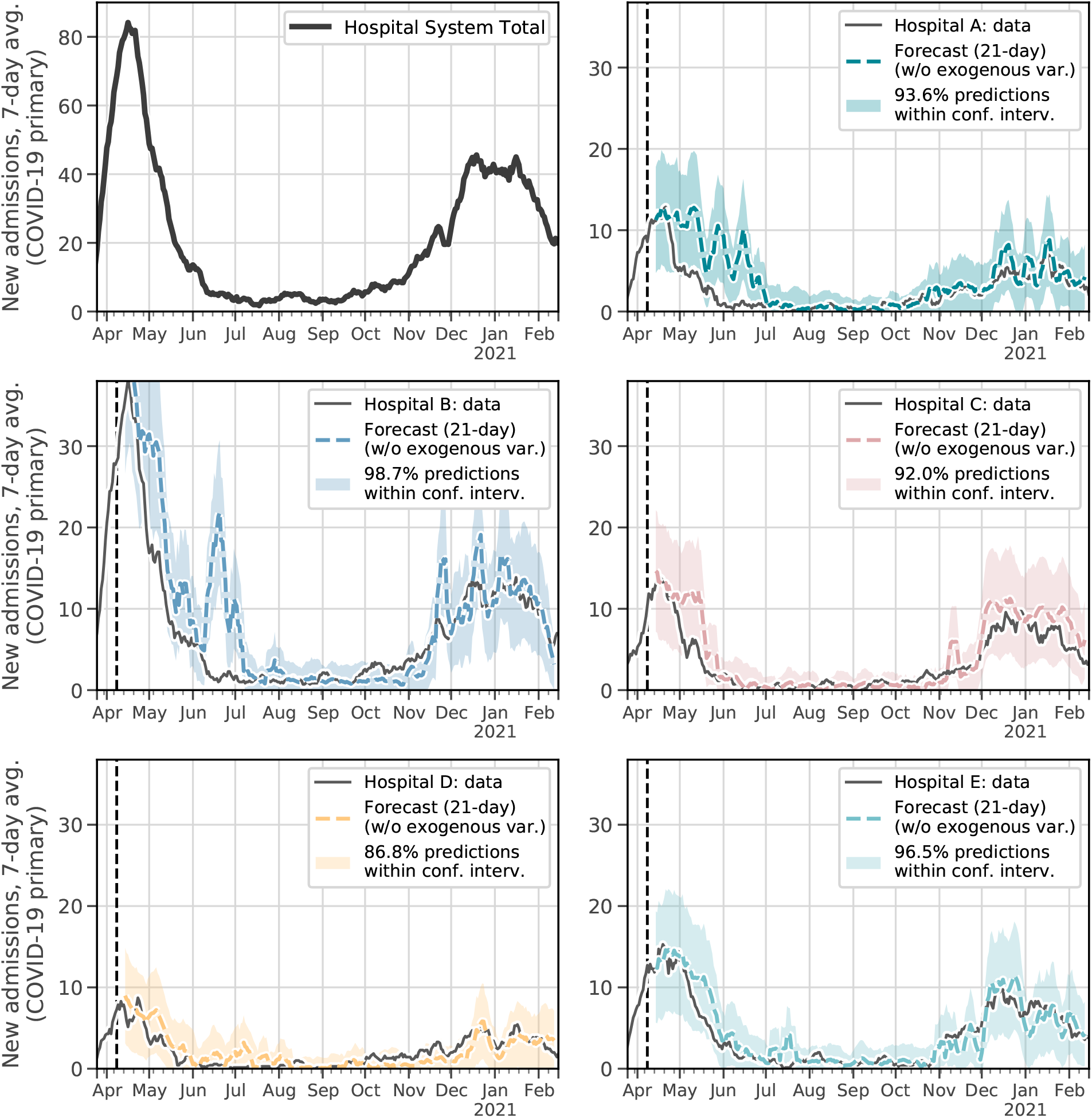
21-day forecast for each hospital without including additional exogenous data. Similar to Fig. 2, the vertical dashed line marks the boundary of boundary of initial training data and future data; visualized at a 7-day rolling average; 95% confidence intervals are shown for each forecast. Hyper-parameters of each model were tuned separately to make sure fair comparisons were made between models.

## Notes

### Author Declarations

In addition to time series data of statewide test positivity percentages from the Massachusetts Department of Public Health, two other data sources were used in this research: aggregated location data from mobile devices and hospital admission data. Because both data sources were only used in aggregated form, the Institutional Review Board (IRB) at Northeastern University determined that the study does not qualify as human subject research and is exempt from IRB approval.

